# Exploring the potential of Claude 2 for risk of bias assessment: Using a large language model to assess randomized controlled trials with RoB 2

**DOI:** 10.1101/2024.07.16.24310483

**Authors:** Angelika Eisele-Metzger, Judith-Lisa Lieberum, Markus Toews, Waldemar Siemens, Felix Heilmeyer, Christian Haverkamp, Daniel Boehringer, Joerg J Meerpohl

## Abstract

Systematic reviews are essential for evidence based healthcare, but conducting them is time and resource consuming. To date, efforts have been made to accelerate and (semi-) automate various steps of systematic reviews through the use of artificial intelligence and the emergence of large language models (LLMs) promises further opportunities. One crucial but complex task within systematic review conduct is assessing the risk of bias of included studies. Therefore, the aim of this study was to test the LLM Claude 2 for risk of bias assessment of 100 randomized controlled trials using the revised Cochrane risk of bias tool (“RoB 2”; involving judgements for five specific domains and an overall judgement). We assessed the agreement of risk of bias judgements by Claude with human judgements published in Cochrane Reviews. The observed agreement between Claude and Cochrane authors ranged from 41% for the overall judgement to 71% for domain 4 (“outcome measurement”). Cohen’s κ was lowest for domain 5 (“selective reporting”; 0.10 (95% confidence interval (CI): −0.10-0.31)) and highest for domain 3 (“missing data”; 0.31 (95% CI: 0.10-0.52)), indicating slight to fair agreement. Fair agreement was found for the overall judgement (Cohen’s κ: 0.22 (95% CI: 0.06-0.38)). Sensitivity analyses using alternative prompting techniques or the more recent version Claude 3 did not result in substantial changes. Currently, Claude’s RoB 2 judgements cannot replace human risk of bias assessment. However, the potential of LLMs to support risk of bias assessment should be further explored.

## INTRODUCTION

Systematic reviews are considered a highly valuable tool for evidence synthesis and informed decision making in healthcare and other fields, however, require a lot of time and resources (1, 2). Steps in conducting a systematic review include framing the research question, preparation of a review protocol, searching for and selecting relevant studies, risk of bias (RoB) assessment of the studies included, data extraction, synthesis and interpretation of the results and finally reporting (3, 4).

In order to make this work more time- and resource efficient, efforts have been underway for several years to assist or even (semi-)automate steps of the systematic review process, using artificial intelligence (AI) and, more specifically, machine learning (ML) techniques (5, 6). Based on (un-/semi-/self-supervised or reinforcement) learning from data provided and further development of pattern recognition systems, algorithms allow to constantly improve performance on specific tasks without being explicitly programmed to do so (7, 8). Exemplary applications that use ML to support steps of systematic reviews include Rayyan (9), Covidence (10) and EPPI Reviewer (11), that are particularly useful to support screening and data extraction, deduplication tools such as Deduklick (12), and the RobotReviewer (13) for RoB assessment.

Recently, further AI systems based on large language models (LLMs) such as ChatGPT (14), PaLM 2 (15), LLaMA (16), or Claude (17) have gained attention, and a variety of potential uses in health care and research alike has been discussed (18–21). LLMs are trained on a very large dataset to always predict the most likely next token, given any textual input. They are commonly fine-tuned to simulate or participate in human dialogues (22). Contrariwise to conventional statistical classification methods, which rely on task specific training using labelled training data, LLMs can be instructed to perform any task without task-specific training. The training process is replaced with crafting and refining detailed instructions in natural language, a process known as prompt-engineering. Limitations of LLMs include the lack of full control including unexpected responses that may contain toxic language, discrimination, or even false (’made up’) information (22–24). So far, a number of attempts to use LLMs for systematic review support have been made, e.g. to help formulating a structured review question (25), screening (26), producing an R code for conducting a meta-analysis (25) or data extraction (27). First experiences are still clearly flawed, albeit promising.

Assessing the RoB in each study included is a pivotal step of a systematic review. For assessing randomized controlled trials (RCTs), the revised Cochrane Risk of Bias tool (“RoB 2”) (28) is considered the gold standard. The tool is structured into five bias domains (1. bias arising from the randomization process, 2. bias due to deviations from intended interventions, 3. bias due to missing outcome data, 4. bias in measurement of the outcome, 5. bias in selection of the reported result). An overall judgement is made on the basis of assessments of each individual domain, each in the categories of “low risk”, “some concerns” or “high risk” (28, 29). RoB assessment not only requires time and at least two reviewers, but also underlies to a degree of subjectivity even when utilizing standardized tools (30–32). Therefore, the objective and reproducible automation of this systematic review step appears particularly important and valuable. Currently, there are very limited methods to support RoB assessment using ML (5). However, also using ChatGPT alone for RoB assessments seems not recommendable, neither for RCTs (33, 34) nor for non-randomized studies of interventions (35), due to limited agreement in RoB judgements between ChatGPT and humans.

Claude 2, first released by Anthropic in March 2023 (17), appears particularly suitable for conducting RoB assessments, perhaps better than ChatGPT: Characterized by a particularly large context window, substantial volumes of data such as full texts of study reports can be processed in one piece – as stated by Anthropic – with a comparatively low rate of hallucinations, high accuracy and robustness (17, 36, 37), making it a promising candidate for supporting RoB assessment. Most recently, in May 2024, Lai et al. (38) first described assessing RoB in RCTs with both ChatGPT and Claude and found substantial accuracy and consistency, however, restricted to a modified version of the original Cochrane RoB-tool (“RoB 1”) from 2011 (39). This tool has been revised in 2019 (28) in order to address some of its limitations and the use of the former tool is no longer recommended (29). Therefore, we aimed at using the revised RoB 2 tool for our study.

In this proof-of concept study, our aim was to determine how well the LLM Claude 2 assesses the RoB of RCTs using the RoB 2 tool compared to conventional RoB 2 assessments published by human reviewers in Cochrane Reviews.

## METHODS

The protocol for this proof-of-concept study has been registered on Open Science Framework (OSF) (https://osf.io/42dnb) on September 11, 2023. We applied a validation study design to evaluate the performance of Claude 2 compared to humans (reference standard).

### Sample and Eligibility Criteria

To identify a sample of recent Cochrane Reviews of interventions applying RoB 2, we searched the Cochrane Library in October 2023 using the search string “ROB2” OR “ROB-2” OR “ROB 2.0” OR “revised cochrane risk-of-bias” (all text) with a limit for publication date from January 2019 onwards and a filter for review type “intervention”. We manually checked each Cochrane Review retrieved and excluded Cochrane Reviews exclusively using RoB assessment tools other than RoB 2. A random sample of 100 2-arm parallel group RCTs was drawn (see sample size estimation), choosing at least one RCT per Cochrane Review. We excluded Cluster-RCTs and cross-over RCTs because RoB assessment methods slightly differ for those types of RCTs. Furthermore, we excluded RCTs published in languages other than English and RCTs published earlier than 2013 due to our assumption that Claude 2 can best process English texts and that the reporting quality of scientific articles has improved in recent years. As Cochrane Reviews often include RoB assessments for more than one outcome and comparison, we selected the RoB assessment for the first listed outcome and first comparison. If the first comparison and first listed outcome did not contain a suitable RCT, we switched to the next outcome/comparison, and so forth.

### Data collection

For each of the selected RCTs, we manually extracted the following data: bibliographic reference details, the results of the RoB assessment of the Cochrane authors (i.e. the judgement for each RoB 2 domain, the overall assessment as well as all text that was provided to support RoB judgements), study location, condition/disease studied, type of intervention (i.e. pharmacological intervention; surgical intervention; non-pharmacological, non-invasive intervention), type of control intervention (i.e. placebo, treatment as usual/other intervention, no intervention), outcome and comparison named in the Cochrane Review (for which RoB was assessed for the selected RCT), original outcome named in the RCT and references to published study protocols and register entries.

### Prompt engineering and generation of Claude RoB assessments

We used Claude 2 (17) to create new RoB assessments for each of the selected RCTs. The testing was performed in February 2024.

#### Prompt engineering

A prompt is an input, usually in textual form, to which the LLM produces an output (40). Prompt engineering refers to the process of developing the most suitable prompt to successfully accomplish a task (41). If a prompt contains one or more variables that are replaced by media (e.g. text extracted from a PDF file), it is referred to as prompt template (40).

During a pilot phase, we developed and refined various prompt templates using different prompting techniques and tested them with a sample of 30 RCTs from three Cochrane Reviews (42–44). These were then excluded from any further analysis or testing. This preliminary testing resulted in one final main prompt template. Two alternative prompt templates that also showed acceptable results during the pilot phase were used for sensitivity testing. All three prompt templates were uploaded on OSF in advance to conducting the actual testing and can be accessed via https://osf.io/2phyt (prompt number 12 is the final main prompt template).

#### Contents of the final main prompt template

Our prompt template asked Claude 2 to assess the RoB of the respective RCT, considering each of the five domains of the RoB 2 tool and to provide an overall judgement. It also specified the format of the judgement options (i.e. “low risk”, “some concerns” or “high risk”) and asked Claude to provide justifying text for each judgement, embedded in a machine readable JSON structure.

The prompt template included the text extracted from the PDF article of the RCT (but no possibly existing additional reports on the same study), the compressed study protocol/analysis plan, if available, or (if no published study protocol/analysis plan was available) the study register entry (e.g. record from https://clinicaltrials.gov), if available. We used the ConvertAPI service (https://www.convertapi.com) to extract the full text of the PDF articles. As the RoB 2 tool is applied specifically per outcome, we also specified the individual outcome for which the assessment should be made (including time of measurement, if more than one follow-up time point was available). These data were injected into the prompt template in an automated fashion using custom software (see below).

We suspected that some of the Cochrane Reviews used for our dataset might have been in the training data for Claude. To avoid a simple recall of the results from the training data, we opted for a full instruction prompt template that does not mention the RoB 2 tool by name, but instead provides a detailed instruction on how to perform the assessment. The instructions were taken from the official RoB 2 full guidance document (45). The RoB 2 tool provides the option to choose between assessing the effect of assignment to intervention (“intention-to-treat” effect) and assessing the effect of adhering to intervention (“per-protocol” effect) for the second domain (RoB due to deviations from the intended interventions). As the first option is usually used for efficacy studies, we only provided guidance for this first method to Claude.

During the pilot phase, we learned that it is helpful to generate separate prompts for each of the RoB 2 domains in order to minimize the reasoning complexity. We concatenated all five LLM responses (one response for each RoB 2 domain) and proceeded with the prompt for overall assessment on this basis. Furthermore, we learned during the pilot phase that the RCT protocols (and register entries) need to be compressed with a separate prompt and injected into the final prompt template, as they can be very lengthy, often longer than the manuscript itself. Assembling the single prompts via manual copy-pasting would have been unfeasible and error-prone. Therefore, we developed a program to automate the process (see below).

#### Program

We used a custom program called “Patchbay” to automate the process of assembling the single prompts, including compression of the RCT protocols, and combined all the necessary components into the final prompts according to the defined templates. This allowed us to efficiently create the number of prompts required for the study while minimizing the risk of errors. The source code and documentation for Patchbay are available at https://github.com/daboe01/LLMPatchbay.

#### Iterations

When using Claude, users can set the temperature, i.e. the randomness of the answers one receives from Claude (36). Lower temperatures lead to more stable and conservative outputs corresponding to the most likely variants while higher temperatures produce more creative and diverse responses (36). For our study, we set the temperature as low as possible. We then performed three iterations, i.e. we ran the prompt template three times for each RCT. This method has recently been used to quantify the uncertainty of LLM outputs (46, 47). If the judgments of the three iterations matched, we selected one at random for our testing (because the justifying text could still vary to some extent). If the judgments did not match, we randomly selected from the results that were more frequent (e.g. if the prompt resulted in one “low risk” judgement and two “some concerns” judgements in a domain, we randomly selected one of the “some concerns” judgements). In the rare cases where all three iterations differed in their assessment, we also selected one at random. This technique is known as “self consistency” (48).

### Data analysis

We quantitatively compared the RoB judgements created by Claude to the judgements of the Cochrane authors (reference standard). For each of the 100 RCTs, judgements of either “low risk”, “some concerns” or “high risk” were available for the five RoB 2 domains as well as the overall assessment. We calculated the performance of Claude using Cohen’s weighted kappa coefficient (κ) for ordinal data (R package ‘psych’) (49–51), a measure of interrater agreement that controls for agreement by chance and can take values between −1.0 to 1.0. We adjusted each Cohen’s κ for clustering in case of more than one RCT per Cochrane Review using the design effect as suggested in the Cochrane Handbook (52, 53). Cohen’s κ was interpreted as poor (<0.00), slight (0.00-0.20), fair (0.21-0.40), moderate (0.41-0.60), substantial (0.61-0.80) and almost perfect (0.81-1.00), as suggested by Landis and Koch (54). Additionally, we calculated the observed percentage of agreement between Claude and the reference standard, sensitivity and specificity as well as the positive and negative predictive value (PPV and NPV) of Claude for i) a high RoB rating (versus “some concerns” or “low risk”) or ii) a low RoB rating (versus “some concerns” or “high risk”) compared to the reference standard. Estimates are given with their 95% confidence intervals (CIs). The R code for calculating the primary results of the manuscript can be accessed at https://osf.io/2phyt.

To identify reasons for non-agreement between Claude and the reference standard, we manually checked justifications provided by Claude and the Cochrane authors with the deviating judgements for the RoB 2 domains 1-5. We reviewed all “two-level-discrepancies” (i.e. “high risk” versus “low risk”, which we regarded as more severe) by comparing given justifications to the content of the original reports and protocols/register entries of the trials. We documented whether we agreed with either the Cochrane authors or Claude or whether we would suggest a “some concerns” judgement instead. Additionally, we reviewed a random sample of 10 discrepancies for each of the five specific RoB 2 domains for the remaining discrepancies “some concerns” versus “low risk” or “some concerns” versus “high risk”. We compared justifications and summarized observed reasons for non-agreement (without comparing them to the original reports of the RCTs, for reasons of feasibility). The overall judgement strongly depends on the judgements for the five specific domains (e.g., to reach an overall low RoB, the study must be judged to be at low RoB for all five domains (45)). Therefore, we checked the 100 overall judgements of Claude for compliance with the algorithm provided in the RoB 2 guidance (45).

#### Additional analyses

We conducted exploratory sensitivity and subgroup analyses as described below. We did not perform any inductive statistics, i.e. the analyses were descriptive only. They had not been pre-specified in our protocol.

##### Sensitivity analyses

To explore the impact of the prompt characteristics on the results, we performed sensitivity analyses, i.e. we repeated the testing for the same 100 RCTs, using two alternative prompt templates. The first alternative prompt template (“step-by-step prompt”) was very similar to the final main prompt template but additionally based on the framework of zero-shot chain of thought prompting (40). The other alternative prompt template (“minimal prompt”) was much shorter, included only very little information taken from the RoB 2 guidance, and is therefore possibly prone to bias from dataset contamination.

Few days after our testing, a new version of Claude was launched (37). We therefore decided to perform an additional sensitivity analysis using Claude 3 Opus and the prompt template that had proven to be most promising in the previous testing. This was conducted in March 2024. We did not perform further prompt engineering using the new version of Claude.

##### Subgroup analyses

We carried out the following subgroup analyses using our final main prompt template:

1. Individual analyses for the different types of interventions studied in the RCTs (due to the low number of surgical interventions, we only performed analyses for pharmacological versus other - non-pharmacological, non-surgical - interventions).
2. Individual analyses according to whether a published study protocol or a register entry was available. We differentiated between RCTs without protocol or register entry and RCTs with at least one (protocol or register) entry.
3. Individual analyses according to whether the three iterations of Claude produced the same results or whether results differed between the three iterations. The rationale for this was that we assumed higher uncertainty and possibly poorer accuracy in the assessments where the iterations differed.

#### Sample size estimation

We assumed a Cohen’s weighted κ of 0.7 (indicating substantial agreement) with a corresponding 95% CI of 0.55-0.85 for the overall RoB rating between Claude and the reference standard. Furthermore, we anticipated proportions of 0.20, 0.50, 0.30 for frequencies of the rating categories (“low risk”, “some concerns”, “high risk”) and an alpha-level of 0.05 (55). This resulted in a minimum of 88 required RCTs for this study. To safely meet these assumptions, we included a sample of 100 RCTs.

## RESULTS

### Sample

Our search for Cochrane Reviews resulted in 78 Cochrane Reviews of interventions fulfilling our eligibility criteria. The search and selection process is illustrated in a PRISMA flow chart (see figure 1). The full sample of Cochrane Reviews assessed for eligibility along with reasons for exclusion is part of the data stored at OSF and can be accessed via https://osf.io/2phyt.

**Figure 1.**
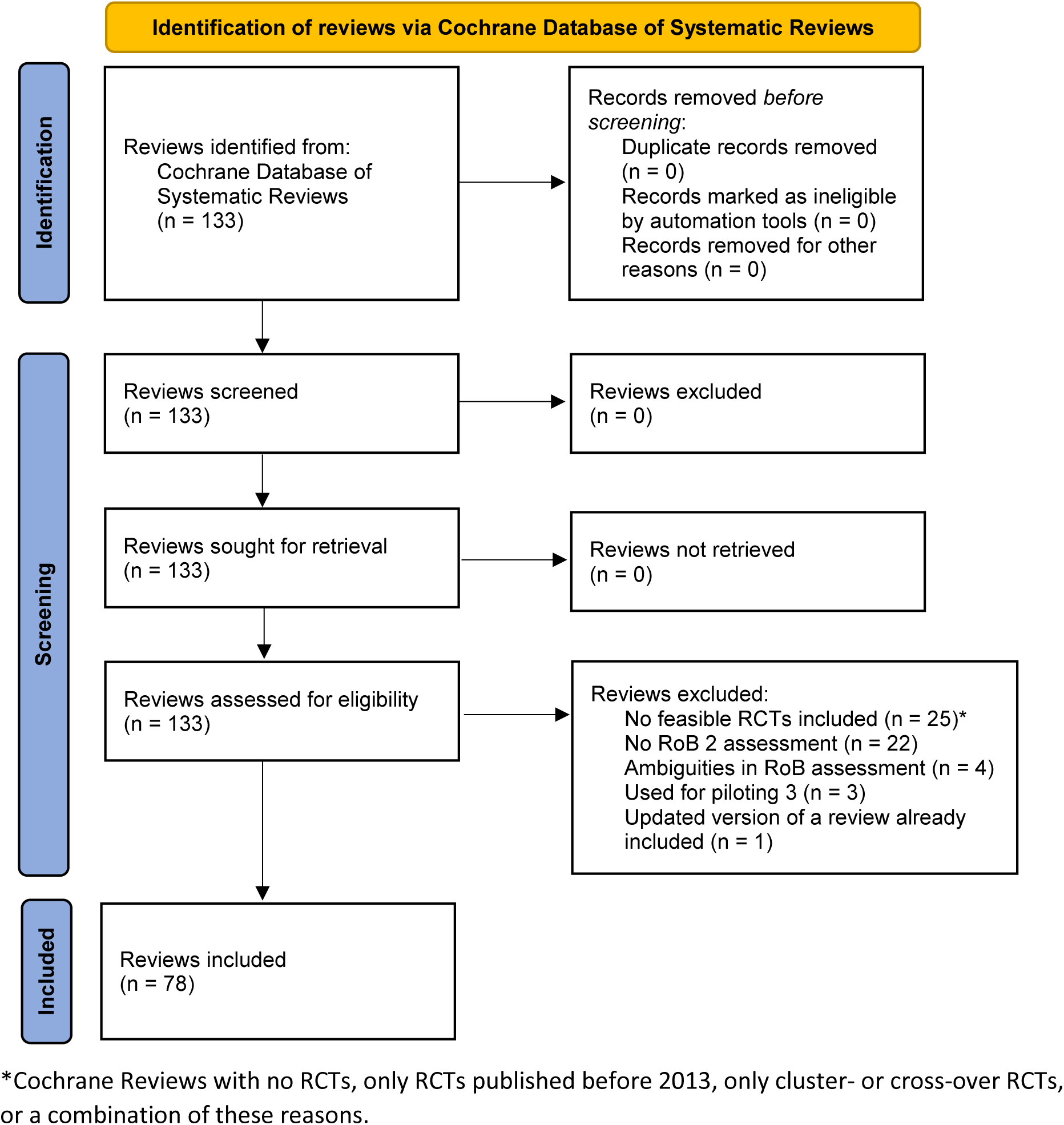
PRISMA flow chart illustrating the search process for Cochrane Reviews of interventions.

Our final sample of 100 RCTs consisted of 56 RCTs drawn from 56 unique Cochrane Reviews and 44 RCTs drawn from a total of 22 Cochrane Reviews (2 per review).

### Study characteristics

The RCTs were published between 2013 and 2022. Fifty RCTs studied non-pharmacological, non-surgical interventions, such as psychological interventions or exercise interventions. Pharmacological interventions were studied in 44 RCTs and surgical interventions in 6 RCTs. The most common condition studied was COVID-19 (18 RCTs). For 32 RCTs, we were able to identify a published study protocol, and for 82, a register entry was available. For 16 RCTs, neither a protocol nor a register entry was available.

Full extracted data with reference to the corresponding Cochrane Reviews and including the full results of our testing can be accessed via https://osf.io/2phyt.

### RoB assessment with Claude

RoB judgements of Claude 2 for the five domains and the overall judgement are summarized in table 1, along with the human judgements of the Cochrane authors (reference standard). The most frequent judgement of Claude 2 for domain 1 to 5 was “low risk” while it judged the overall domain most frequently with “some concerns”. “High risk” judgements occurred rarely. We had no missing values but Claude occasionally deviated from the prescribed response format (e.g. returned “unable to assess” or “no information”; this occurred three times in the results of the final main prompt template). As we performed three iterations (see methods) per RCT, we finally received at least one valid judgement.

**Table 1.**
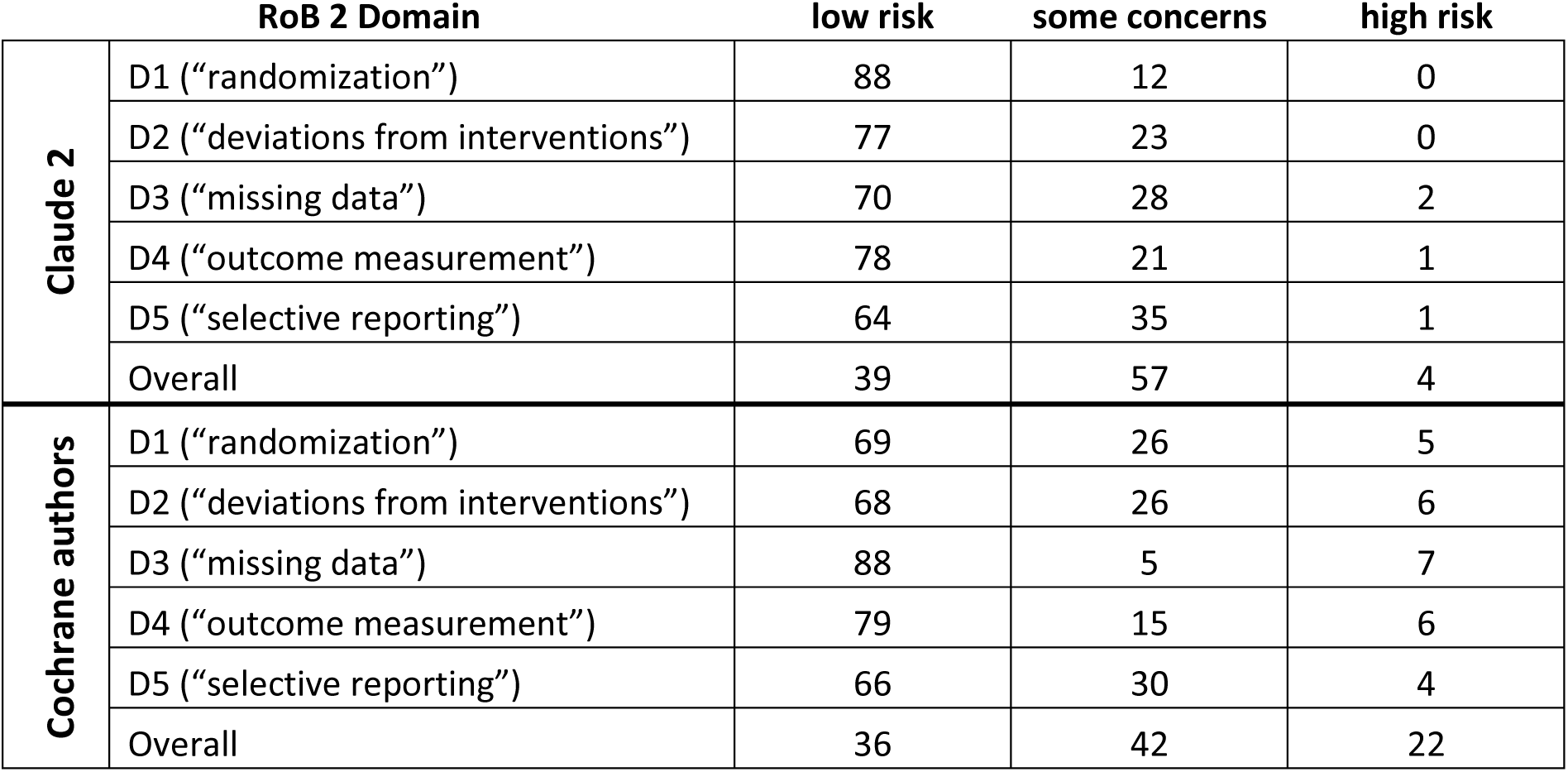
Risk of bias judgements of Claude 2 and Cochrane authors (number of judgements per RoB 2 domain, n=100 RCTs).

| RoB 2 Domain |  | low risk | some concerns | high risk |
| --- | --- | --- | --- | --- |
| Claude 2 | D1 ("randomization") | 88 | 12 | 0 |
|  | D2 ("deviations from interventions") | 77 | 23 | 0 |
|  | D3 ("missing data") | 70 | 28 | 2 |
|  | D4 ("outcome measurement") | 78 | 21 | 1 |
|  | D5 ("selective reporting") | 64 | 35 | 1 |
|  | Overall | 39 | 57 | 4 |
| Cochrane authors | D1 ("randomization") | 69 | 26 | 5 |
|  | D2 ("deviations from interventions") | 68 | 26 | 6 |
|  | D3 ("missing data") | 88 | 5 | 7 |
|  | D4 ("outcome measurement") | 79 | 15 | 6 |
|  | D5 ("selective reporting") | 66 | 30 | 4 |
|  | Overall | 36 | 42 | 22 |

Table 2 shows the overall judgements of Claude 2 tabulated against the overall judgements of the Cochrane authors. Tables for the remaining 5 RoB 2 domains can be found in the supplement (table S1-S5). Figure 2 illustrates the overall RoB judgements of Claude versus Cochrane authors using a Sankey diagram (56, 57).

**Figure 2.**
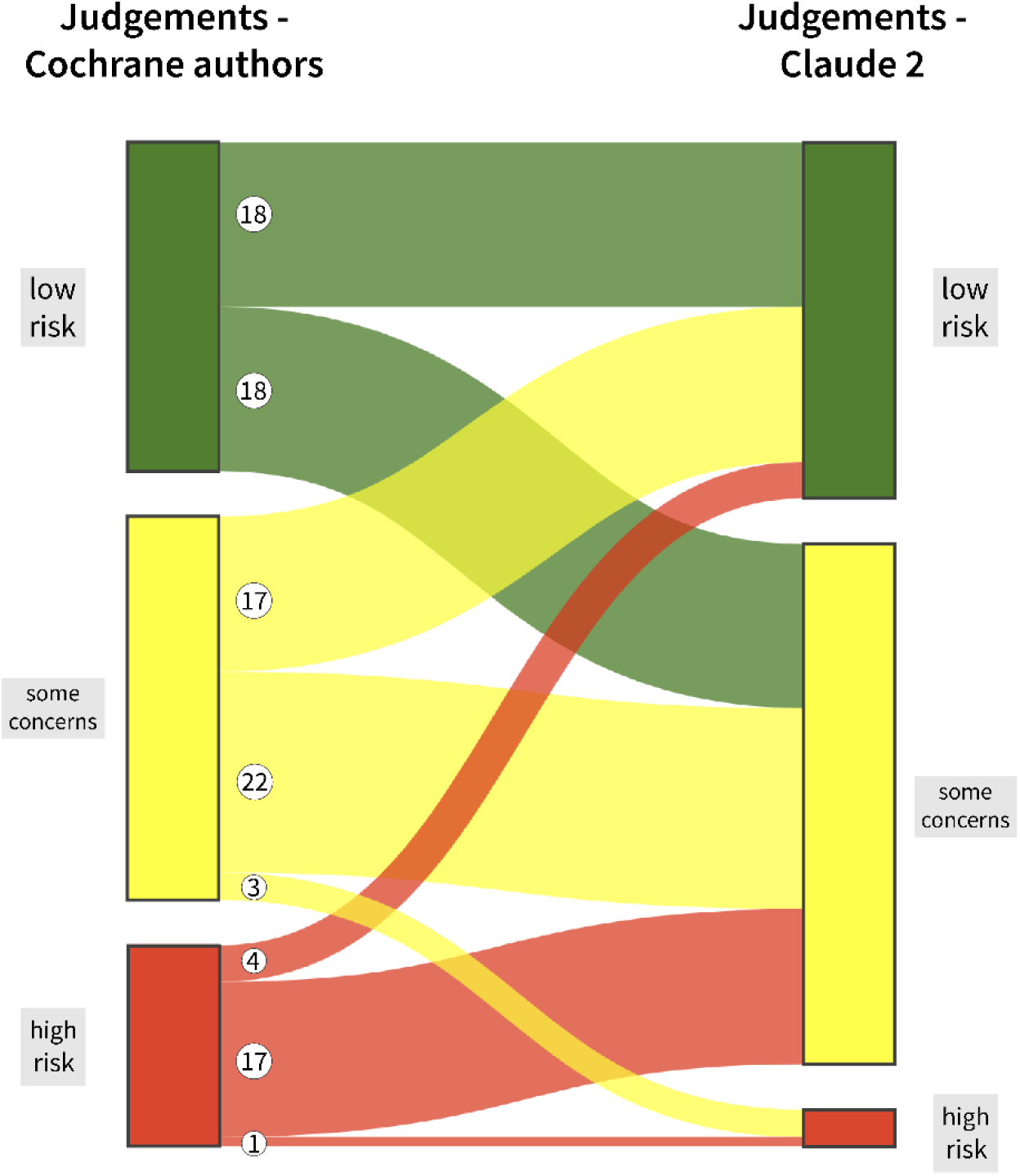
Sankey diagram illustrating differing and congruent overall risk of bias judgements of the Cochrane authors and Claude 2. An animated version of this figure can be accessed via https://osf.io/2phyt.

**Table 2.** Overall risk of bias judgements of Claude 2 tabulated against the overall judgements of the Cochrane authors (n=100 RCTs).

|  |  | Cochrane Review |  |  |  |
| --- | --- | --- | --- | --- | --- |
|  |  | low risk | some concerns | high risk | Total |
| Claude 2 | low risk | 18 | 17 | 4 | 39 |
|  | some concerns | 18 | 22 | 17 | 57 |
|  | high risk | 0 | 3 | 1 | 4 |
|  | Total | 36 | 42 | 22 | 100 |

The observed percentage of agreement, Cohen’s weighted κ, sensitivity, specificity and predictive values are displayed in table 3. Given the low number of “high risk” judgements of Claude, we only present sensitivity, specificity, PPV and NPV of Claude for a low RoB rating (versus “some concerns” and “high risk”). Values for a high RoB rating (versus “low risk” and “some concerns”) are presented in the supplement (table S6).

**Table 3.** Performance of Claude 2 compared to the Cochrane authors (n=100 RCTs).

| RoB 2 Domain | Agreement | Cohen's Kappa<br>(95% CI) | Sensitivity<br>(95% CI) | Specificity<br>(95% CI) | PPV<br>(95% CI) | NPV<br>(95% CI) |
| --- | --- | --- | --- | --- | --- | --- |
| D1 ("randomization") | 65% | 0.11 (-0.08; 0.29) | 0.90 (0.80; 0.96) | 0.16 (0.05; 0.34) | 0.70 (0.67; 0.74) | 0.42 (0.20; 0.67) |
| D2 ("deviations from interventions") | 63% | 0.12 (-0.08; 0.32) | 0.81 (0.70; 0.89) | 0.31 (0.16; 0.50) | 0.71 (0.66; 0.76) | 0.43 (0.27; 0.61) |
| D3 ("missing data") | 70% | 0.31 (0.10; 0.52) | 0.76 (0.66; 0.85) | 0.75 (0.43; 0.95) | 0.96 (0.89; 0.98) | 0.30 (0.21; 0.41) |
| D4 ("outcome measurement") | 71% | 0.15 (-0.11; 0.41) | 0.81 (0.71; 0.89) | 0.33 (0.15; 0.57) | 0.82 (0.77; 0.86) | 0.32 (0.18; 0.50) |
| D5 ("selective reporting") | 58% | 0.10 (-0.10; 0.31) | 0.68 (0.56; 0.79) | 0.44 (0.27; 0.62) | 0.70 (0.63; 0.77) | 0.42 (0.30; 0.55) |
| Overall | 41% | 0.22 (0.06; 0.38) | 0.50 (0.33; 0.67) | 0.67 (0.54; 0.78) | 0.46 (0.35; 0.58) | 0.70 (0.62; 0.78) |

CI: confidence interval; PPV: positive predictive value; NPV: negative predictive value.
Sensitivity, specificity, PPV and NPV for a low RoB rating versus "some concerns" and "high risk".
Interpretation notes:
Sensitivity (true positive rate): proportion correctly classified as "low risk" by Claude in relation to all "low risk" judgements by the Cochrane authors (reference standard)
Specificity (true negative rate): proportion correctly classified as "some concerns" or "high risk" by Claude in relation to all "some concerns" or "high risk" judgements by the Cochrane authors (reference standard)
PPV: proportion correctly classified as "low risk" by Claude in relation to all "low risk" judgements by Claude (index test)
NPV: proportion correctly classified as "some concerns" or "high risk" by Claude in relation to all "some concerns" or "high risk" judgements by Claude (index test).

The observed agreement between judgements of Claude and judgements of the Cochrane authors ranged from 41% for the overall judgement to 71% for domain 4 (“outcome measurement”). Cohen’s κ was lowest for domain 5 (“selective reporting”; 0.10 [95% CI: −0.10-0.31]) and highest for domain 3 (“missing data”; 0.31 [95% CI: 0.10-0.52]), indicating slight to fair agreement. For the overall judgement, Cohen’s κ was 0.22 (95% CI: 0.06-0.38) which can be interpreted as “fair”. There was strong variation for sensitivity (range 0.50 [95% CI: 0.33-0.67] to 0.90 [95% CI: 0.80-0.96]), specificity (range 0.16 [95% CI: 0.05-0.34] to 0.75 [95% CI: 0.43-0.95]), PPV (range 0.46 [95% CI: 0.35-0.58] to 0.96 [95% CI: 0.89-0.98]) and NPV (range 0.30 [95% CI: 0.21-0.41] to 0.70 [95% CI: 0.62-0.78]). Of note, the width of the confidence intervals (including much lower or higher values) must be considered when interpreting these values.

#### Reasons for non-agreement

##### Review of two-level-discrepancies

We identified 18 two-level-discrepancies (i.e. “low risk” versus “high risk” judgements) for the 5 specific RoB 2 domains: three for D1, four for D2, three for D3, five for D4 and three for D5. All but two of these 18 discrepancies comprised a “high risk” judgement of the Cochrane authors and a “low risk judgements of Claude 2. For 12 judgements, we would have opted for a “some concerns” judgement instead of the differing judgements of Claude and the Cochrane authors, and for six judgements, we agreed with the decisions of the Cochrane authors. There was no case in which we agreed with Claude’s judgement. Two examples of two-level-discrepancies between Claude and the Cochrane authors are provided in table 4 with additional comments.

**Table 4.** Examples for two-level-discrepancies between Claude and reference standard, with comments and suggested judgement by the authors of this article.

| RoB 2 domain | Reference<br>Cochrane<br>Review, RCT | Judgement and justifying text of the Cochrane authors | Judgement and justifying text<br>of Claude 2 | Comment of the authors of this<br>article; suggested judgement |
| --- | --- | --- | --- | --- |
| D1 („randomization“) | Richardson,<br>Dale (58),<br>Hung, Chow<br>(59) | **High risk**<br>The allocation sequence was probably random, but it seems likely that the research assistants who enrolled participants may have been able to foresee allocation: “The first author generated a random allocation sequence table. Two research assistants were responsible for then enrolling and assigning the participants to either the experimental group or a waitlist control group”. Baseline differences did not suggest a problem with the randomisation process. | **Low risk**<br>The allocation sequence was random and baseline differences between groups were compatible with chance, suggesting adequate randomization. | We agree with the Cochrane authors that the allocation sequence seems to not have been concealed; however, as there are no significant baseline differences, this would result in a “**some concerns**” judgement, following the algorithm in the RoB 2 guidance. |
| D3 („missing data“) | Schrijver,<br>Lenferink<br>(60), Jolly,<br>Sidhu (61) | **High risk**<br>“There was imbalance in the follow-up rates between telephone health coaching (82.7%; 37 withdrawals) and usual care (96.2%; 7 withdrawals).” p. 4.<br>“Participants who did not provide data at 12 months were more likely to be in GOLD stage 3, to be smokers, had lower levels of self-reported physical activity, and to live alone than responders.” p. 5.<br>Comment: For health-related quality of life, data were available for 75% of intervention patients (217 of 289), although some sensitivity analyses were performed. | **Low risk**<br>The percentage of missing outcome data is below 5% and the missing data likely did not bias the results. Therefore, there is low risk of bias due to missing outcome data. | We disagree with Claude that the percentage of missing data is below 5%. The numbers cited by the Cochrane authors seem to be correct. Numbers of participants lost to follow-up differed between intervention groups; reasons for missing outcome data were not reported; therefore we think that giving “**high risk**” is reasonable. |

##### Review of other discrepancies

Below, we give the main identified reasons for disagreement (“some concerns” versus either “low risk” or “high risk”) between Claude and the Cochrane authors for each domain of the RoB 2 tool.

- *D1 (“randomization“):* One main reason for discrepancies in this domain was that Claude assumed appropriate concealment of allocation while the Cochrane authors criticized lacking (information on) allocation concealment.
- *D2 (“deviations from interventions“):* Differences in dealing with lack of blinding (of participants or carers) were one main reason for discrepant judgements. E.g., Claude judged “some concerns” in some cases where Cochrane authors regarded it as unlikely that deviations from the intended interventions had occurred due to the open-label design and judged “low risk”.
- *D3 (“missing data“):* Reasons for discrepancies comprised different interpretations of the potential influence of missing data (i.e. Claude regarded the amount of missing data as less or more concerning, compared to the Cochrane authors), but Claude also seemed to have overseen data in some cases (e.g. reported different percentage of missing data, compared to the Cochrane authors).
- *D4 (“outcome measurement“):* For this domain, justifications especially deviated regarding information on assessor blinding (e.g. Claude assumed assessors to be blinded while the Cochrane authors stated they were aware of the allocated intervention) and the impact of non-blinded assessors on the validity of outcome assessment.
- *D5 (“selective reporting“):* One main reason for discrepancies in this domain was that Claude either ignored the absence of a pre-specified protocol/analysis plan or did not consider an available protocol.
- *Overall judgement:* Of the 100 available overall judgements of Claude, only 2 clearly deviated from the algorithm provided in the RoB 2 guidance (45), i.e. Claude judged the overall RoB as “low”, but assessed single domains as “some concerns”.

#### Results of the additional analyses

The observed percentage of agreement and Cohen’s κ values for the sensitivity and subgroup analyses are given in the supplement (table S7-S11). These analyses were descriptive only.

##### Sensitivity analyses

Below, we summarize the results of the sensitivity analyses using two alternative prompt templates and using the latest version of Claude (Claude 3). Cohen’s κ values obtained in the sensitivity analyses indicate slight to fair agreement between reference standard and Claude for the RoB judgements, with two exceptions for RoB 2 domains that had values >0.40, indicating moderate agreement (i.e. domain 4 “outcome measurement” using Claude 2 with the “step-by-step” prompt template and domain 1 “randomization” using Claude 3 with the “step-by-step” prompt template).

- “*Step-by-step” prompt template*: The observed agreement values were comparable to the values obtained using the final main prompt template (42% agreement for the overall judgement). Cohen’s κ had a slightly wider range (0.08 [95% CI: −0.13-0.28] to 0.43 [95% CI: 0.20-0.66], highest κ for domain 4 “outcome measurement”) and was 0.28 (95% CI: 0.11-0.46) for the overall judgement.
- *“Minimal” prompt template*: This prompt resulted in slightly higher observed agreement for all domains (47% for the overall judgement) and a slightly larger range for Cohen’s κ (−0.04 [95% CI: 0.12-0.04] to 0.40 [95% CI: 0.19-0.61], highest κ for domain 1 “randomization”) with a lower Cohen’s κ for the overall judgement (0.19 [95% CI: 0.00-0.38]), when compared to the values obtained using the final main prompt template.
- *“Step-by-step” prompt template with Claude 3*: Generally, the observed agreement and Cohen’s κ were comparable to the other runs using Claude 2, with some variation, but with no apparent pattern. We obtained 45% observed agreement and a Cohen’s κ of 0.19 (95% CI: 0.02-0.37) for the overall judgement. Cohen’s κ values had a larger range compared to the runs with Claude 2 (0.08 [95% CI: −0.07-0.23] to 0.54 [95% CI: 0.36-0.72], highest κ for domain 1 “randomization”).

##### Subgroup analyses

Below, we summarize the results of the subgroup analyses for RCTs on pharmacological versus other (non-pharmacological, non-surgical) interventions, RCTs without protocol/register entry versus RCTs with at least one of protocol/register entry and RCTs for which the three iterations of Claude produced the same results versus differing results.

- *Pharmacological (n=44) versus other (n=50) interventions:* Cohen’s κ values for all domains were slightly lower for RCTs on pharmacological interventions (range −0.11 [95% CI: −0.36-0.15] to 0.27 [95% CI: 0.01-0.53], highest κ for domain 3 “missing data”) compared to RCTs on other interventions (range 0.11 [95% CI: −0.12-0.35] to 0.36 [95% CI: 0.05-0.66], highest κ for domain 3 “missing data”). The observed agreement for the overall judgement was 38.6% for pharmacological interventions and 42% for other interventions.
- *No protocol/register entry (n=16) versus at least one of protocol/register entry (n=83)*: All in all, the observed agreement and Cohen’s κ values were comparable for the two groups, with some variation but no striking differences, except for domain 3 (“missing data”). Cohen’s κ for this domain was 0.41 (95% CI: 0.01-0.82) for the group of RCTs without protocol/register entry, compared to 0.28 (95% CI: 0.06-0.51) for the group of RCTs with at least one of protocol or register entry.
- *For the three iterations of Claude: Same results (n=68) versus differing results (n=32) of the iterations:* The range of observed agreement and Cohen’s κ values was comparable for the two groups. One notable difference was that Cohen’s κ for the group of RCTs with differing results for the three iterations was highest (0.32; 95% CI: 0.11-0.53) for the overall judgement, which was not the case in any other analysis.

## DISCUSSION

In this work, we compared RoB assessments of RCTs created by the LLM Claude 2 with assessments created by human reviewers and published in Cochrane Reviews. To our knowledge, this is the first study that uses Claude to assess RCTs applying the RoB 2 tool. We found only slight to fair agreement between Claude and humans for all RoB domains when using our final main prompt template. Only in the sensitivity analyses, using two alternative prompting approaches, we obtained moderate agreement for two domains, i.e. domain 4 “outcome measurement” and domain 1 “randomization”. Based on these results, we infer that Claude should currently not be used as a stand-alone tool to conduct RoB assessment of included studies within the systematic review process.

Additional sensitivity and subgroup analyses did not indicate that our results differed substantially depending on specific characteristics. Thus, it did, e.g., not seem to make a great difference whether a protocol or register entry was available or whether the trial was on pharmacological or other interventions. Using alternative prompt templates or the novel version of Claude also did not substantially change our results.

Reasons for disagreement between Claude and the Cochrane authors include, e.g., that possible problematic features of the trials (such as lack of blinding of participants, carers or assessors or a certain proportion of missing data) were assessed differently. In some cases, Claude also provided wrong information in the supporting text or obviously missed details. Among the 18 two-level-discrepancies (“low risk” versus “high risks”), which we verified by consulting the original articles, there were 12 cases for which we would have opted for a “some concerns” judgement instead of the judgements made by Claude and the Cochrane authors. This highlights that judgements made using the RoB 2 tool underlie a certain degree of subjectivity.

Indeed, also agreement of RoB 2 judgements between humans is far from perfect (62, 63). Additionally, adherence of systematic reviewers to RoB 2 guidance is often poor (64). In a study by Minozzi and colleagues (62), four raters independently used the RoB 2 tool to assess RoB for 70 outcomes of 70 RCTs on various unrelated topics and obtained only slight agreement (Fleiss’ κ of 0.16) for the overall assessment. This is even lower than the agreement between Claude and the Cochrane authors we obtained for the overall assessment in our study. In a follow-up study by Minozzi and colleagues (63), four raters independently applied RoB 2 for 80 results related to seven outcomes reported in 16 RCTs on a similar topic. During a pilot run of the tool (“calibration exercise”), they developed an implementation document specific for this topic in advance. They were then able to increase their interrater agreement from no agreement (Fleiss’ κ of −0.15) during the calibration exercise to finally moderate agreement (Fleiss’ κ of 0.42) for the overall assessment. This implies that, in addition to using the RoB 2 guidance, further consultations and agreements, related to the specific topic of interest for a systematic review, might be necessary to increase reliability of RoB 2 assessments. Thus, comparing RoB 2 assessments by Claude to this “imperfect” and variable reference standard obviously is problematic.

Just recently, other authors have used LLM to conduct RoB assessment, with mixed results. Pitre et al. (34) found comparably low agreement between ChatGPT-4 and Cochrane authors when assessing RoB of 157 RCTs from 34 Cochrane Reviews using RoB 2 (Cohen’s κ of 0.16 for the overall assessment). Testing the use of ChatGPT (GPT-4) for RoB assessment of non-randomized studies of intervention using ROBINS-I (65), Hasan et al. (35) also obtained only slight agreement (Cohen’s κ of 0.13 for the overall assessment). In contrast, Lai et al. (38) reported promising results when using ChatGPT and Claude (versions not specified) to assess RoB of 30 RCTs from three systematic reviews using a modified version of the original Cochrane RoB tool (“RoB 1”) (39): In their study, Cohen’s κ ranged from 0.54 to 0.96 for ChatGPT and from 0.76 to 0.96 for Claude for the different domains (there is no overall judgement included in RoB 1). Although there were some important differences in methodology, such as using another tool that is obviously easier to apply, using only 30 RCTs on only three different topics and calculating agreement from only 2 possible judgements (i.e. “low risk” or “high risk”), their results still seem surprising. Therefore, there is a need to further explore LLM support for RoB assessment of research studies. Future studies should probably also focus on LLM support going beyond the production of stand-alone RoB judgements, for example automatic extraction of the relevant content of an RCT that needs to be reviewed to assess its RoB.

### Strengths and limitations

We used a large sample of RCTs drawn from the largest possible number of Cochrane Reviews on various topics for our study. Additionally, we used a thoroughly elaborated prompting approach and also explored two alternative prompt templates. Nevertheless, our work has a number of limitations. First, reproducibility of our testing is limited due to the variations of LLMs in producing output. As development of LLMs is progressing, it is likely that the reproducibility of our results decreases further in the future. Secondly, as pointed out above, we had to compare RoB 2 judgements of Claude to an “imperfect” human reference standard, for which we know that it is variable and interrater agreement is poor. However, as the “true” RoB 2 assessments are unknown, using assessments from different Cochrane authors was, perhaps, the most appropriate method to obtain a reference standard. RoB 2 is currently the recommended tool to assess RoB in RCTs, making its use indispensable. Lastly, Claude had only access to the main article and the compressed protocol or (if no protocol was available) register entry. We did not provide Claude with any supplementary material or further articles on the same study, while the Cochrane authors presumably consulted as many sources as were necessary and available. Compressing the protocols and register entries using an extra prompt was necessary due to their often extensive length.

## Conclusion

Based on our results, the use of Claude to conduct RoB assessment of RCTs using RoB 2 is currently not recommended. Further investigation is needed to explore LLM support for RoB assessment of research studies. Other models of use than providing stand-alone RoB judgements could additionally be investigated. In conclusion, RoB assessment of RCTs included in high-quality systematic reviews currently still requires at least two independent human reviewers.

## Supporting information

Supplement_Eisele-Metzger2024

## ACKNOWLEDGEMENTS

We would like to thank Kathrin Grummich for her support in developing the search strategy, Philipp Kapp for his advice on the use of RoB 2 and Georg Melber for his assistance in the visualization of data.

## DATA AVAILABILITY STATEMENT

Prompt templates, the R code used for analysis, model responses, extracted data and the full sample of Cochrane Reviews assessed for eligibility are stored at open science framework (OSF) and can be accessed via the following link: https://osf.io/2phyt. The source code and documentation for our custom program (Patchbay) are available at https://github.com/daboe01/LLMPatchbay.

## ETHICS STATEMENT

We used secondary data published in Cochrane Reviews and RCT reports. Ethical approval was therefore not necessary.

