## Supplement_Eisele-Metzger2024 for "Exploring the potential of Claude 2 for risk of bias assessment: Using a large language model to assess randomized controlled trials with RoB 2"

**Table S1**. Risk of bias judgements of Claude 2 for the RoB 2 domain 1 (“randomization”) tabulated against the judgements of the Cochrane authors (n=100 RCTs).

| | | **Cochrane Review** | | | |
|---|---|---|---|---|---|
| | | **low risk** | **some concerns** | **high risk** | **Total** |
| **Claude 2** | **low risk** | 62 | 23 | 3 | 88 |
| | **some concerns** | 7 | 3 | 2 | 12 |
| | **high risk** | 0 | 0 | 0 | 0 |
| | **Total** | 69 | 26 | 5 | 100 |

**Table S2**. Risk of bias judgements of Claude 2 for the RoB 2 domain 2 (“deviations from interventions”) tabulated against the judgements of the Cochrane authors (n=100 RCTs).

| | | **Cochrane Review** | | | |
|---|---|---|---|---|---|
| | | **low risk** | **some concerns** | **high risk** | **Total** |
| **Claude 2** | **low risk** | 55 | 18 | 4 | 77 |
| | **some concerns** | 13 | 8 | 2 | 23 |
| | **high risk** | 0 | 0 | 0 | 0 |
| | **Total** | 68 | 26 | 6 | 100 |

**Table S3**. Risk of bias judgements of Claude 2 for the RoB 2 domain 3 (“missing data”) tabulated against the judgements of the Cochrane authors (n=100 RCTs).

| | | **Cochrane Review** | | | |
|---|---|---|---|---|---|
| | | **low risk** | **some concerns** | **high risk** | **Total** |
| **Claude 2** | **low risk** | 67 | 1 | 2 | 70 |
| | **some concerns** | 20 | 3 | 5 | 28 |
| | **high risk** | 1 | 1 | 0 | 2 |
| | **Total** | 88 | 5 | 7 | 100 |

**Table S4**. Risk of bias judgements of Claude 2 for the RoB 2 domain 4 (“outcome measurement”) tabulated against the judgements of the Cochrane authors (n=100 RCTs).

| | | **Cochrane Review** | | | |
|---|---|---|---|---|---|
| | | **low risk** | **some concerns** | **high risk** | **Total** |
| **Claude 2** | **low risk** | 64 | 9 | 5 | 78 |
| | **some concerns** | 15 | 6 | 0 | 21 |
| | **high risk** | 0 | 0 | 1 | 1 |
| | **Total** | 79 | 15 | 6 | 100 |

**Table S5**. Risk of bias judgements of Claude 2 for the RoB 2 domain 5 (“selective reporting”) tabulated against the judgements of the Cochrane authors (n=100 RCTs).

| | | Cochrane Review | | | |
|---|---|---|---|---|---|
| | | **low risk** | **some concerns** | **high risk** | **Total** |
| **Claude 2** | **low risk** | 45 | 17 | 2 | 64 |
| | **some concerns** | 20 | 13 | 2 | 35 |
| | **high risk** | 1 | 0 | 0 | 1 |
| | **Total** | 66 | 30 | 4 | 100 |

**Table S6**. Accuracy values for the performance of Claude 2 compared to the Cochrane authors (for “high risk” versus “some concerns” or “low risk”; n=100 RCTs).

| **RoB 2 Domain** | **Sensitivity (95% CI)** | **Specificity (95% CI)** | **PPV (95% CI)** | **NPV (95% CI)** |
|---|---|---|---|---|
| D1 (“randomization”) | n.a. | n.a. | n.a. | n.a. |
| D2 (“deviations from interventions”) | n.a. | n.a. | n.a. | n.a. |
| D3 (“missing data”) | 0.00 (0.00; 0.41) | 0.98 (0.92; 1.00) | 0.23 (0.06; 0.62) | 0.94 (0.92; 0.95) |
| D4 (“outcome measurement”) | 0.17 (0.00; 0.64) | 1.00 (0.96; 1.00) | 0.55 (0.15; 0.84) | 0.95 (0.94; 0.97) |
| D5 (“selective reporting”) | 0.00 (0.00; 0.60) | 0.99 (0.94; 1.00) | 0.26 (0.06; 0.65) | 0.97 (0.95; 0.98) |
| Overall | 0.05 (0.00; 0.23) | 0.96 (0.89; 0.99) | 0.25 (0.04; 0.75) | 0.78 (0.76; 0.80) |

CI: confidence interval; PPV: positive predictive value; NPV: negative predictive value; n.a.: not applicable (no ratings “high risk” for D1 and D2, therefore not calculable).

**SENSITIVITY ANALYSES**

**Table S7**. Performance of Claude 2 (using the "step-by-step prompt" and the "minimal" prompt) compared to the Cochrane authors (n=100 RCTs).

| RoB 2 Domain | „Step-by-step" Prompt | | „Minimal" Prompt | |
|---|---|---|---|---|
| | **Agreement** | **Cohen's Kappa (95% CI)** | **Agreement** | **Cohen's Kappa (95% CI)** |
| D1 ("randomization") | 69% | 0.18 (-0.01; 0.37) | 73% | 0.40 (0.19; 0.61) |
| D2 ("deviations from interventions") | 53% | 0.08 (-0.13; 0.28) | 64% | -0.04 (-0.16; 0.07) |
| D3 ("missing data") | 67% | 0.30 (0.09; 0.52) | 78% | 0.33 (0.11; 0.55) |
| D4 ("outcome measurement") | 77% | 0.43 (0.20; 0.66) | 73% | 0.32 (0.09; 0.54) |
| D5 ("selective reporting") | 62% | 0.11 (-0.09; 0.30) | 63% | -0.04 (-0.12; 0.04) |
| Overall | 42% | 0.28 (0.11; 0.46) | 47% | 0.19 (0.00; 0.38) |

CI: confidence interval.

**Table S8**. Performance of Claude 3 (using the "step-by-step prompt") compared to the Cochrane authors (n=100 RCTs).

| RoB 2 Domain | Agreement | Cohen's Kappa (95% CI) |
|---|---|---|
| D1 ("randomization") | 73% | 0.54 (0.36; 0.72) |
| D2 ("deviations from interventions") | 41% | 0.08 (-0.07; 0.23) |
| D3 ("missing data") | 55% | 0.28 (0.11; 0.44) |
| D4 ("outcome measurement") | 59% | 0.28 (0.12; 0.44) |
| D5 ("selective reporting") | 58% | 0.22 (0.03; 0.41) |
| Overall | 45% | 0.19 (0.02; 0.37) |

CI: confidence interval.

**SUBGROUP ANALYSES**

**Table S9**. For RCTs on pharmacological versus other (non-pharmacological, non-surgical) interventions: Performance of Claude 2 compared to the Cochrane authors.

| RoB 2 Domain | **Pharmacological interventions** (n=44 RCTs) | | **Other interventions** (n=50 RCTs) | |
|---|---|---|---|---|
| | **Agreement** | **Cohen's Kappa (95% CI)** | **Agreement** | **Cohen's Kappa (95% CI)** |
| D1 ("randomization") | 75.0% | 0.05 (-0.20; 0.31) | 56.0% | 0.11 (-0.12; 0.35) |
| D2 ("deviations from interventions") | 59.1% | 0.08 (-0.22; 0.38) | 62.0% | 0.11 (-0.17; 0.39) |
| D3 ("missing data") | 68.2% | 0.27 (0.01; 0.53) | 70.0% | 0.36 (0.05; 0.66) |
| D4 ("outcome measurement") | 77.3% | -0.01 (-0.22; 0.19) | 68.0% | 0.21 (-0.16; 0.58) |
| D5 ("selective reporting") | 47.7% | -0.11 (-0.36; 0.15) | 64.0% | 0.30 (0.01; 0.59) |
| Overall | 38.6% | 0.10 (-0.13; 0.34) | 42.0% | 0.30 (0.08; 0.52) |

CI: confidence interval; RCTs: randomized controlled trials; 6 RCTs not included in this analysis studied surgical interventions.

**Table S10**. For RCTs with versus without study protocol or register entry: Performance of Claude 2 compared to the Cochrane authors.

| RoB 2 Domain | **Protocol/register entry available** (n=83 RCTs) | | **Neither protocol nor register entry available** (n=16 RCTs) | |
|---|---|---|---|---|
| | **Agreement** | **Cohen's Kappa (95% CI)** | **Agreement** | **Cohen's Kappa (95% CI)** |
| D1 ("randomization") | 68.7% | 0.14 (-0.11; 0.40) | 50.0% | 0.07 (-0.08; 0.21) |
| D2 ("deviations from interventions") | 62.7% | 0.15 (-0.08; 0.39) | 62.5% | 0.00 (0.00; 0.00) |
| D3 ("missing data") | 69.9% | 0.28 (0.06; 0.51) | 68.8% | 0.41 (0.01; 0.82) |
| D4 ("outcome measurement") | 73.5% | 0.18 (-0.14; 0.50) | 56.3% | 0.06 (-0.25; 0.37) |
| D5 ("selective reporting") | 59.0% | 0.01 (-0.21; 0.23) | 56.3% | 0.20 (-0.20; 0.60) |
| Overall | 39.8% | 0.22 (0.03; 0.40) | 43.8% | 0.21 (-0.08; 0.49) |

CI: confidence interval; RCTs: randomized controlled trials; for one RCT without register entry not included in this analysis it was deemed unclear whether a protocol was available.

**Table S11**. For RCTs where the three iterations of Claude produced the same results versus differing results: Performance of Claude 2 compared to the Cochrane authors.

| RoB 2 Domain | **Same results for the 3 iterations** (n= 68 RCTs) | | **Differing results for the 3 iterations** (n= 32 RCTs) | |
|---|---|---|---|---|
| | **Agreement** | **Cohen‘s Kappa (95% CI)** | **Agreement** | **Cohen‘s Kappa (95% CI)** |
| D1 (“randomization”) | 63.2% | 0.02 (-0.15; 0.19) | 68.8% | 0.27 (-0.09; 0.63) |
| D2 (“deviations from interventions”) | 67.6% | 0.08 (-0.10; 0.26) | 53.1% | 0.16 (-0.18; 0.50) |
| D3 (“missing data”) | 75.0% | 0.37 (0.11; 0.63) | 59.4% | 0.22 (-0.08; 0.53) |
| D4 (“outcome measurement”) | 76.5% | 0.11 (-0.10; 0.32) | 59.4% | 0.17 (-0.32; 0.65) |
| D5 (“selective reporting”) | 57.4% | 0.12 (-0.12; 0.36) | 59.4% | 0.04 (-0.31; 0.38) |
| Overall | 42.6% | 0.15 (-0.04; 0.34) | 37.5% | 0.32 (0.11; 0.53) |

CI: confidence interval; RCTs: randomized controlled trials.
